# Agreement between self-reported COVID-19 and dried blood spot serology

**DOI:** 10.1101/2025.04.08.25325501

**Authors:** Nicola Sheppard, Matthew TC Carroll, Brigitte M Borg, Zheng Quan Toh, Paul V Licciardi, Catherine L. Smith, Jillian F. Ikin, Michael J. Abramson, Karen Walker-Bone, Tyler J Lane

**Affiliations:** School of Public Health and Preventive Medicine, Monash University, Melbourne, Australia; Monash Rural Health Churchill, Monash University, Churchill, Australia; Respiratory Medicine, Alfred Health, Melbourne, Australia; Infection, Immunity and Global Health Theme, Murdoch Children’s Research Institute, Parkville, Australia; Department of Paediatrics, University of Melbourne, Parkville, Australia

## Abstract

**Introduction:** Identifying likely COVID-19 cases with good accuracy is essential for epidemiological research on the pandemic’s health effects. Therefore, evaluating case detection methods for cost-effectiveness and reliability is important. We investigated the agreement between a validated self-report questionnaire for COVID-19 and dried blood spot serology for SARS-CoV-2 antibodies.

**Methods:** Between June and October 2023, 311 adults completed the self-report COVID-19 questionnaire validated by the Avon Longitudinal Study of Parents and Children, and provided fingertip blood samples which underwent Enzyme Linked Immunosorbent Assay to quantify IgG antibodies to SARS-CoV-2 nucleocapsid (N)-protein. We applied several statistical approaches to assess agreement: Cohen’s Kappa measure of inter-rater reliability; positive (PPV) and negative (NPV) predictive values; logistic regressions of the year of most recent self-reported COVID-19 on serostatus; and linear regressions of N-protein antibody concentrations.

**Results:** Two-thirds (203, 65%) of participants self-reported a history of COVID-19 whereas one-third (98, 32%) were seropositive for SARS-CoV-2 N-protein antibodies. Across all years, there was only “fair” agreement (κ = 0.23 [95%CI 0.15 - 0.31]). Self-reported COVID-19 had a PPV of 41% and an NPV of 87% for SARS-CoV-2 seropositivity. PPV was low for 2020-21 (36%) and 2022 (33%), but higher (75%) for participants whose most recent case was in 2023. Compared to participants with no self-reported history of COVID-19, those reporting SARS-CoV-2 infection in 2023 had 23 times greater odds of being seropositive (95%CI: 9.18-62.5), and had 1,079% (617-1,838%) higher N-protein concentrations, after adjustment for confounders.

**Conclusion:** A validated COVID-19 self-report questionnaire is useful for identifying people who have previously had COVID-19 and may be reasonably accurate in identifying when an infection occurred, at least within a calendar year. Furthermore, consistent with existing evidence, serological testing becomes much less sensitive over time.

## Introduction

In the five years since its emergence, COVID-19 has attracted an unprecedented volume of research (1). The urgency of sharing knowledge about COVID-19, particularly during the early days of the pandemic, led to research being published faster than ever before (2), igniting concerns about its quality (1,3,4). One of the most important public health metrics pertaining to COVID-19 has been case numbers. Such data are vital to better understand SARS-CoV-2 infectivity and inform public health policy. Evaluating methods of COVID-19 case detection remains important to understanding the reliability of data and to finding cost-effective ways of monitoring the ongoing impact of COVID-19.

Polymerase Chain Reaction (PCR) tests, which detect viral RNA in swabs of fluid taken from the nose and mouth during acute infection are widely accepted to be the reference standard for COVID-19 diagnosis (4,5). However, PCR sensitivity wanes over a few weeks (6), thus missing those who cannot access testing during acute infection. Furthermore, PCR testing programs that target symptomatic individuals miss between 32-47% of cases who do not exhibit symptoms (8-10). Rapid Antigen Tests (RATs), which use a similar sampling method to PCR but detect viral proteins instead of RNA, can also be used at the time of acute infection. However, these have similar shortcomings to PCR testing and poorer sensitivity (7).

Serology testing, which identifies viral antibodies in a blood sample, can retrospectively identify SARS-CoV-2 infections. In contrast to PCR sensitivity, which wanes in the weeks following infection, the diagnostic accuracy of serology testing increases over the same period (6). However, detectable COVID-19 antibody levels in the blood also eventually wane over the months following infection, particularly in those with milder disease (8–11). Detectable antibody levels also vary depending on the assay used (12,13).

Self-report questionnaires are another means to retrospectively capture COVID-19 infection. However, these too can yield a high rate of false positives as well as false negatives due to asymptomatic cases (14–17). Alternatively, questionnaires that ask individuals to self-report prior COVID-19 diagnoses by PCR or medical opinion proved to be a valuable resource during the pandemic (18).

Our study evaluated the agreement between two methods of retrospectively detecting COVID-19: a serology test measuring SARS-CoV-2 antibodies in dried blood spot (DBS) samples (19) and a validated self-report questionnaire for COVID-19 (20).

## Methods

### Population

Participants were recruited from the Hazelwood Health Study (HHS) Adult Cohort, which was established to investigate the long-term health effects of the 2014 Hazelwood open cut coal mine fire in the Latrobe Valley, southeastern Australia (21).The present study took place among Cohort members in the third round of the Respiratory Stream (22) and the long-term respiratory follow-up (23), who agreed to provide a DBS sample and answer questions about their COVID-19 history during clinical visits between June and October 2023.

### COVID-19 indicators

We used two measures of determining prior SARS-CoV-2 infection based on data collected during the 2023 clinical visit: a validated self-report questionnaire and a serology test performed on a DBS sample.

The self-report COVID-19 questionnaire was validated by the Avon Longitudinal Study of Parents and Children (ALSPAC) (20). It asked, “Do you think that you currently have or have had COVID-19?”, and had four potential responses: 1) Yes, confirmed by a positive test; 2) Yes, suspected by a doctor but not tested; 3) Yes, my own suspicions;4) No.” We also asked participants about the year of their COVID-19 infection(s), allowing for repeat infections. Participants who answered “Yes” to any of questions 1-3 were classified as having self-reported COVID-19. “Yes” respondents were subcategorised into the year of their most recent reported infection.

The selected serology test was an Enzyme Linked Immunosorbent Assay (ELISA) which quantified IgG antibodies to SARS-CoV-2 nucleocapsid (N)-protein in the blood. The ELISA was adapted from a previously published Spike-protein IgG assay and the cutoff value for seropositivity was chosen based on testing in pre-pandemic samples (24). ELISA Units per millilitre (EU/mL) ≥30.94 was categorised as seropositive, while <21.52 EU/mL was seronegative. Values between 21.52 to 30.93 EU/mL were classified as “equivocal” and treated as seronegative for the purposes of the main analysis.Participants whose titre was too high to read were recoded to 1050 EU/mL. For sensitivity analyses, equivocal cases were treated as seropositive.

Antibody levels were measured in DBS samples. A few drops of blood from the participant’s finger were placed on a Guthrie Card, which was then dried, sealed and stored at room temperature before being transported to the Murdoch Children’s Research Institute in Melbourne for analysis. This approach has several advantages over serology testing using venous blood samples: DBS samples are less invasive, require less expert skill to collect and are easier to store and transport than vials of blood. Serological results using DBS samples strongly correlate with serology results from venepuncture using the selected assay (19).

### Analysis

We initially characterised the sample using descriptive statistics. Then, we compared the annual prevalence of self-reported COVID-19 in our sample to the annual incidence of cases in the local government areas of Latrobe City and Wellington (from which our sample was drawn), using data from the Victorian Department of Health (25) and Australian Bureau of Statistics (26).

We applied several statistical approaches to assess agreement between serostatus and self-reported COVID-19. We first calculated Cohen’s Kappa, a measure of inter-rater reliability that accounts for the likelihood of agreement by chance, to determine agreement between serostatus and self-reported COVID-19 in any year. Second, we calculated the positive and negative predictive values (PPV and NPV) of self-reported COVID-19 using serostatus as a reference standard, both overall and then by year of most recent self-reported COVID-19. Third, we conducted logistic regressions of the year of most recent self-reported COVID-19 on serostatus, both crude and adjusted for age at clinical visit (natural spline with 3 degrees of freedom to account for non-linear effects), sex, and education level (secondary to year 10, secondary to year 11-12, certificate/trade/university qualification). The fourth approach was to conduct crude and linear regressions of N-protein antibody concentrations, which were log-transformed to present results as proportional change (27). Finally, we performed sensitivity analyses that combined seropositive and equivocal antibody concentrations. Analyses using year of most recent self-reported COVID-19 combined 2020 and 2021 due to small numbers.

### Ethics

This study was approved by the Monash University Human Research Ethics Committee as part of the Hazelwood Health Study: Respiratory Stream Round 3 (Project ID: 36471) and the Alfred Hospital Ethics Committee (project number 90/21). All participants provided written informed consent to participate in this study.

## Results

### Descriptives

Of the 821 members of the Hazelwood Health Study Adult Cohort invited to participate, 318 (39%) attended the clinic, and 312 (38%) provided blood samples for serology. One had unreadable serology data and was therefore excluded from the analysis. Characteristics of the 311 participants included in the analysis are listed in Table 1.

**Table 1.** Participant characteristics.

| Characteristic | Study sample<br>(*n* = 311; *n* (%) or median [IQR]) |
| --- | --- |
| Female | 176 (57%) |
| Age | 63 years [IQR: 52, 71] |
| N-protein antibodies to SARS-CoV-2 | 9 EU/mL [IQR: 5, 43] |
| Seropositive for SARS-CoV-2 antibodies | 98 (32%) |
| *Equivocal* | 14 (4.5%) |
| Any self-reported COVID-19 | 203 (65%) |
| Confirmed by test | 201 (65%) |
| Own suspicions | 2 (0.6%) |
| Most recent year of self-reported COVID-19* |  |
| 2020 | 7 (2.3%) |
| 2021 | 21 (6.8%) |
| 2022 | 134 (43%) |
| 2023 | 40 (13%) |
| Don't know | 1 (<0.1%) |
| Never had COVID-19 | 108 (35%) |

\* Total number of participants reporting COVID-19 in each year (as opposed to most recent only) were as follow: 2020 – 7, 2021 - 33, 2022 - 151, 2023 - 40; self-report and serology data were collected during a 2023 clinical visit; “most recent self-reported COVID-19” refers to retrospective answers of when infection occurred

Two-thirds (203, 65%) of respondents self-reported a history of COVID-19, with all but two reporting that the diagnosis was based on a positive test result. Conversely, one-third (98, 32%) of participants were seropositive for SARS-CoV-2 N-protein antibodies, indicating past infection. By year, most self-reported infections were in 2022, corresponding to the biggest surge in infections observed in Victorian Department of Health data (25) and seropositive blood donors (28).

In Figure S1 (Appendix), we compare study sample annual prevalence to population-level annual incidence in Latrobe City and Wellington local government areas.^1^ Notably, while both have an outsized peak in 2022, the relative magnitude compared to other years was largest in the population Latrobe City and Wellington data. For instance, the 2022 annual incidence in Latrobe City and Wellington was 37.3%, 17.8 times greater than next highest annual incidence year, which was 2.1% in 2023; in contrast, the equivalent figures for annual prevalence in the study sample was 48.6% in 2022 but only 3.7 times greater than 2023 where it was 12.9%.

### Agreement between self-reported COVID-19 and serostatus

Across all years, there was only “fair” agreement (κ = 0.23 [95%CI 0.15 - 0.31]) between self-reported COVID-19 and seropositivity (29). Using serostatus as a reference standard, self-reported COVID-19 had a PPV of 41% and an NPV of 87% for SARS-CoV-2 seropositivity. These results are presented alongside the 2×2 contingency table in Table2. The sensitivity analysis which grouped equivocal with seropositive cases did not meaningfully affect this level of agreement (κ = 0.24 [0.15 - 0.32]; see Table S1).

**Table 2.** 2×2 table of COVID-19 cases based on self-report and serostatus.

|  | Seropositive | Seronegative | Totals | Predictive Agreement |
| --- | --- | --- | --- | --- |
| Self-report + | 84 | 119 | **203** | 41% |
| Self-report - | 14 | 94 | **108** | 87% |
| **Totals** | **98** | **213** | **311** |  |
| **Note: self-report and serology data were collected during a 2023 clinical visit** |  |  |  |  |

### Agreement between self-reported COVID-19 by year and serostatus

Table 3 presents the number of self-reported COVID-19 cases in each year by serostatus, along with the PPV. If a participant had multiple infections, they were counted in the year of their most recent infection. NPV was not calculated for this analysis as ‘most recent self-reported COVID-19’ lacks true negatives (e.g., a participant whose most recent self-reported COVID-19 was in 2023 could also report COVID-19 in a previous year). PPV was low for 2020-21 (36%) and 2022 (33%), but higher (75%) for those whose most recent case was in 2023, (within 9 months of DBS sampling).

**Table 3.** Number of self-reported COVID-19 cases in each year by serostatus, and positive predictive value of the questionnaire using serostatus as a reference standard.

| Most recent self-reported COVID-19 | Seropositive | Seronegative | Positive Predictive Value |
| --- | --- | --- | --- |
| 2020-21 | 10 | 18 | 36% |
| 2022 | 44 | 90 | 33% |
| 2023 | 30 | 10 | 75% |

**Note:** self-report and serology data were collected during a 2023 clinical visit; "most recent self-reported COVID-19" refers to respective answers of when infection occurred

In logistic regression analysis, those with most recent SARS-CoV-2 infection in 2023 had 23 times greater odds of being seropositive (95%CI: 9.18-62.5) compared to those who self-reported no history of COVID-19, after adjusting for potential confounders. Those with self-reported infection in earlier years had lower odds but were still positively associated with seropositive DBS. These results are illustrated in Figure 1 and model results in Table S2. Sensitivity analyses that grouped equivocal with seropositive cases were not meaningfully different (Figure S2 and Table S3).

**Figure 1.**
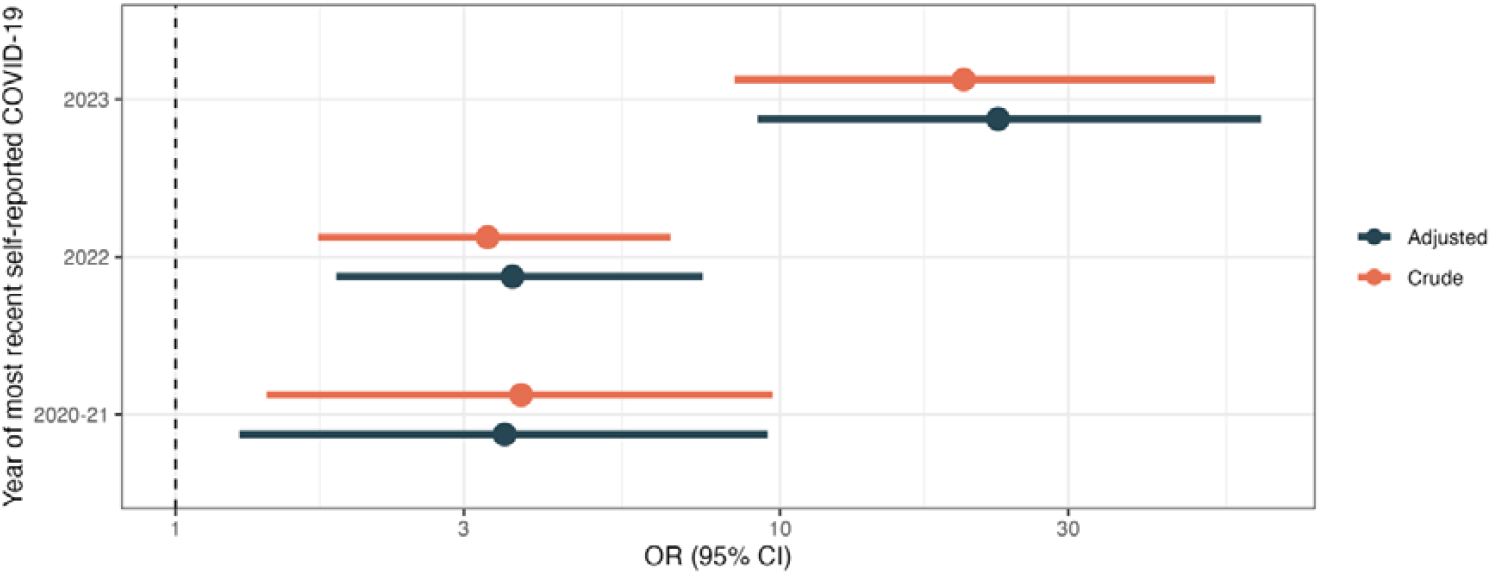
Crude and adjusted (age, sex, educational attainment) odds of seropositive dried blood spot test by year of most recent self-reported COVID-19; self-report and serology data were collected during a 2023 clinical visit; “most recent self-reported COVID-19” refers to retrospective answers of when infection occurred

### Most recent self-reported COVID-19 as predictor of N-protein antibody concentration

Distributions of N-protein concentrations by year of most recent self-reported COVID-19 are illustrated in Figure 2. This shows much wider distributions and higher values in the 2023 group, and clustering in the seronegative category for all groups.

**Figure 2.**
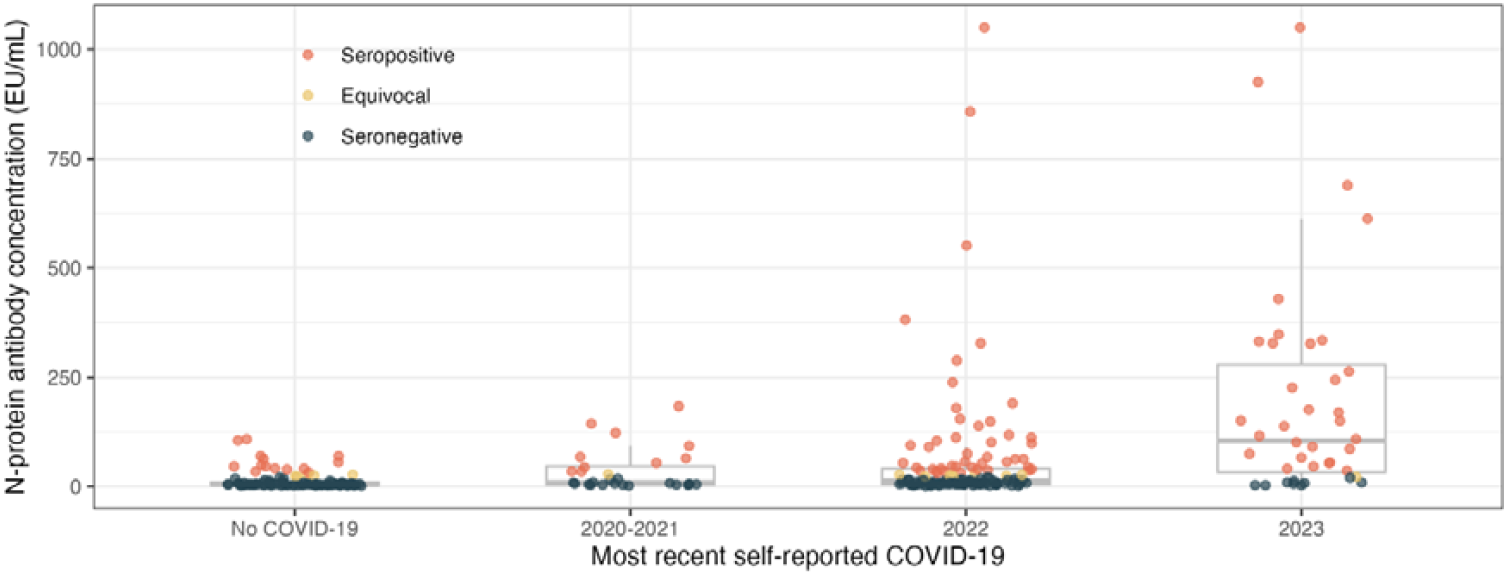
Distributions of antibody concentrations (EU/mL) and status by year of most recent self-reported COVID-19; self-report and serology data were collected during a 2023 clinical visit; “most recent self-reported COVID-19” refers to retrospective answers of when infection occurred

Results of linear regressions are reported in Figure S3 and Table S4. Compared to participants with no self-reported history of COVID-19, those who reported their most-recent case in 2020-2021 had 108% (95%CI: 18-268%) greater N-protein concentrations, those reporting their most recent case in 2022 had 151% (77-255%) greater N-protein concentrations, and those reporting their most recent case in 2023 had 1,079% (617-1,838%) higher N-protein concentrations, after adjustment for confounders.

## Discussion

We found surprisingly weak agreement between self-reported COVID-19 and seropositive DBS. However, there were several interesting findings that could inform others looking for practical and affordable means to identify participants with a history of COVID-19. First, more recent cases of self-reported COVID-19 had greater odds of being seropositive. This aligned with what is known about serum antibodies waning over the months following SARS-CoV-2 infection (8–11). Surprisingly, the NPV of the self-report questionnaire was quite high (87%), indicating those who reported no history of COVID-19 were unlikely to have SARS-CoV-2 antibodies, despite a concern that self-reported COVID-19 would undercount asymptomatic cases. On the other hand, since seropositivity is related to disease severity (11), there may be a gap in our serology data of asymptomatic cases, which could lead to an overestimate of the accuracy of self-reported COVID-19.

Another challenge was relying on participants to accurately recall when they had COVID-19. We added this question on timing of illness to a previous questionnaire (23), but had not evaluated its validity. Our respondents reported a surge in cases in 2022, which was consistent with the recognised surge in cases regionally, likely due to the end of the lockdown era in Victoria combined with the emergence of the highly infectious Omicron variant (30). However, the relative magnitude of this spike compared to other years was considerably larger in the broader population of Latrobe and Wellington local government areas, suggesting some recall bias.

Our findings reflect the messy reality of capturing accurate health data, particularly in the context of COVID-19. But researchers must do their best with resource constraints and available tools and still find meaning to guide policy and practice. One silver lining of this study was the performance of self-reported COVID-19 measures. While self-reported COVID-19 questions are not perfect (particularly for asymptomatic cases) and there was some inaccuracy in participants recalling *when* an infection occurred (cases reported within the same calendar year only achieving 75% PPV with serology), they may nevertheless be a cost-effective way to identify historical cases. This is valuable considering seropositivity wanes rapidly after infection and may render serology tests inaccurate within a few months. This study contributes to understanding of the usefulness of both self-report and serology sampling for COVID-19 disease detection.

### Strengths and limitations

This study is novel in its evaluation of a validated COVID-19 self-report questionnaire in an Australian context. Australia is a high-income country which had state-subsidised PCR testing, thorough contact tracing and relatively low case numbers prior to its vaccination program. Importantly, we were able to adjust our results for confounders such as age, sex and level of education.

A limitation of this study was that neither of the two indicators of COVID-19 that we compared were a true “gold standard”. Furthermore, it is challenging to assess the impact of recall bias on questionnaire results over time, given that the comparator test (DBS serology) also becomes less reliable over time. Lastly, widespread testing in Australia in the early part of the pandemic may have provided participants with more objective data about their history of COVID-19 than was available in countries which did not offer widespread testing, thus limiting this study’s generalisability. While <1% of our sample self-reported COVID-19 based on suspicions alone, the ALSPAC study (based in the UK where infection rates were high before widespread testing or vaccination were available), found that a much larger proportion (12.8%) reported the diagnosis based on suspicion alone (20). Furthermore, these findings may not be generalisable to younger age groups, in whom there tend to be a higher rate of asymptomatic or mild disease, or places where whole-virus vaccines are commonly used as the antibodies they produce are not usually distinguishable from SARS-CoV-2 infection (31).

## Conclusion

Our findings suggest that a validated COVID-19 self-report questionnaire is useful for identifying people who have previously had COVID-19 and may be reasonably accurate in identifying when an infection occurred, at least within a calendar year. Furthermore, consistent with existing evidence, we showed that serological testing becomes much less sensitive over time.

## Data Availability

The study data are confidential and therefore cannot be shared.

## Acknowledgment

The Hazelwood Health Study is funded by the Victorian Department of Health. However, the views in this paper are those of the authors and not the Department. We wish to thank Faizel Hartley, Jacqui Kleiner, Isabella Chicas and Thomas McCrabb for collecting the dried blood spots from participants and William Coote who assisted with fieldwork administration.

## Footnotes

1 While COVID-19 annual incidence (number of cases in a population, which can include multiple infections per individual) and prevalence (persons within a population/sample who have had an infection, where each individual counts only once even if they have multiple infections in the same year) differ, they can be considered roughly comparable, especially as COVID-19 imparts some short-lived immunity to reduce the number of infections within a year.

## References

1. Struelens MJ, Vineis P. COVID-19 Research: Challenges to Interpret Numbers and Propose Solutions. Front Public Health. 2021 Apr 12;9:651089.

2. Horbach SPJM. Pandemic publishing: Medical journals strongly speed up their publication process for COVID-19. Quantitative Science Studies. 2020 Aug;1(3):1056–67.

3. Glasziou PP, Sanders S, Hoffmann T. Waste in covid-19 research. BMJ. 2020 May 12;m1847.

4. Therapeutic Goods Administration (TGA). COVID-19 testing in Australia: information for health professionals | Therapeutic Goods Administration (TGA) [Internet]. Therapeutic Goods Administration (TGA); 2022 [cited 2024 Oct 17]. Available from: https://www.tga.gov.au/products/covid-19/covid-19-tests/covid-19-testing-australia-information-health-professionals

5. CDC. COVID-19. 2024 [cited 2024 Oct 17]. Testing for COVID-19. Available from: https://www.cdc.gov/covid/testing/index.html

6. Miller TE, Garcia Beltran WF, Bard AZ, Gogakos T, Anahtar MN, Astudillo MG, et al. Clinical sensitivity and interpretation of PCR and serological COVID-19 diagnostics for patients presenting to the hospital. The FASEB Journal. 2020;34(10):13877–84.

7. Jegerlehner S, Suter-Riniker F, Jent P, Bittel P, Nagler M. Diagnostic accuracy of a SARS-CoV-2 rapid antigen test in real-life clinical settings. International Journal of Infectious Diseases. 2021 Aug;109:118–22.

8. Fogh K, Larsen TG, Hansen CB, Hasselbalch RB, Eriksen ARR, Bundgaard H, et al. Self-reported long COVID and its association with the presence of SARS-CoV-2 antibodies in a danish cohort up to 12 months after infection. Ramage H, editor. Microbiol Spectr. 2022 Dec 21;10(6):e02537–22.

9. Cromer D, Juno JA, Khoury D, Reynaldi A, Wheatley AK, Kent SJ, et al. Prospects for durable immune control of SARS-CoV-2 and prevention of reinfection. Nat Rev Immunol. 2021 Jun;21(6):395–404.

10. Dan JM, Mateus J, Kato Y, Hastie KM, Yu ED, Faliti CE, et al. Immunological memory to SARS-CoV-2 assessed for up to 8 months after infection. Science. 2021 Feb 5;371(6529):eabf4063.

11. Röltgen K, Powell AE, Wirz OF, Stevens BA, Hogan CA, Najeeb J, et al. Defining the features and duration of antibody responses to SARS-CoV-2 infection associated with disease severity and outcome. Sci Immunol. 2020 Dec 18;5(54):eabe0240.

12. Muecksch F, Wise H, Batchelor B, Squires M, Semple E, Richardson C, et al. Longitudinal serological analysis and neutralizing antibody levels in coronavirus disease 2019 convalescent patients. The Journal of Infectious Diseases. 2021 Feb 13;223(3):389–98.

13. Zheng X, Duan RH, Gong F, Wei X, Dong Y, Chen R, et al. Accuracy of serological tests for COVID-19: A systematic review and meta-analysis. Front Public Health. 2022 Dec 16;10:923525.

14. Terças-Trettel ACP, Muraro AP, Andrade ACDS, Oliveira ECD. Self-reported symptoms and seroprevalence against SARS-CoV-2 in the population of Mato Grosso: a household-based survey in 2020. Rev Assoc Med Bras. 2022 Jul;68(7):928–34.

15. McDonald SA, Miura F, Vos ERA, Van Boven M, De Melker HE, Van Der Klis FRM, et al. Estimating the asymptomatic proportion of SARS-CoV-2 infection in the general population: Analysis of nationwide serosurvey data in the Netherlands. Eur J Epidemiol. 2021 Jul;36(7):735–9.

16. Mulchandani R, Taylor-Philips S, Jones HE, Ades AE, Borrow R, Linley E, et al. Association between self-reported signs and symptoms and SARS-CoV-2 antibody detection in UK key workers. Journal of Infection. 2021 May;82(5):151–61.

17. Piumatti G, Amati R, Richard A, Baysson H, Purgato M, Guessous I, et al. Associations between Depression and Self-Reported COVID-19 Symptoms among Adults: Results from Two Population-Based Seroprevalence Studies in Switzerland. IJERPH. 2022 Dec 12;19(24):16696.

18. Northstone K, Suarez-Perez A, Matthews S, Crawford M, Timpson NJ. The Avon Longitudinal Study of Parents and Children - a resource for COVID-19 research: questionnaire data capture July 2021 to December 2021, with a focus on long COVID [version 2; peer review: 1 approved, 1 approved with reservations]. Wellcome Open Res [Internet]. 2023;8(292). Available from: 10.12688/wellcomeopenres.19596.2

19. Toh ZQ, Higgins RA, Anderson J, Mazarakis N, Do LAH, Rautenbacher K, et al. The use of dried blood spots for the serological evaluation of SARS-CoV-2 antibodies. Journal of Public Health. 2022 Jun 27;44(2):e260–3.

20. Northstone K, Howarth S, Smith D, Bowring C, Wells N, Timpson NJ. The Avon Longitudinal Study of Parents and Children - A resource for COVID-19 research: Questionnaire data capture April-May 2020 [version 2; peer review: 2 approved]. Wellcome Open Res [Internet]. 2020;5(127). Available from: 10.12688/wellcomeopenres.16020.2

21. Ikin J, Carroll MTC, Walker J, Borg B, Brown D, Cope M, et al. Cohort profile: The Hazelwood Health Study Adult Cohort. Int J Epidemiol. 2021 Jan 23;49(6):1777–8.

22. O’Sullivan CF, Smith CL, Gao CX, Borg BM, Lane TJ, Brown D, et al. Echoes from the mine: lung function across a decade following exposure to coal mine fire smoke.

23. Lane TJ, Carroll M, Borg BM, McCaffrey TA, Smith CL, Gao CX, et al. Respiratory symptoms after coalmine fire and pandemic: A longitudinal analysis of the Hazelwood Health Study adult cohort. Babu GR, editor. PLOS Glob Public Health. 2025 Jan 22;5(1):e0004186.

24. Toh ZQ, Anderson J, Mazarakis N, Neeland M, Higgins RA, Rautenbacher K, et al. Comparison of seroconversion in children and adults with mild COVID-19. JAMA Netw Open. 2022 Mar 9;5(3):e221313.

25. Department of Health (Victoria). Victorian Government Data Directory. 2020 [cited 2024 Jul 3]. All Victorian SARS-CoV-2 cases by local government area and postcode 25-Jan-2020 to 14-Sep-2023. Available from: https://discover.data.vic.gov.au/dataset/all-victorian-sars-cov-2-cases-by-local-government-area-and-postcode

26. Australian Bureau of Statistics. Regional population, 2022-23 financial year [Internet]. 2024 [cited 2024 Oct 21]. Available from: https://www.abs.gov.au/statistics/people/population/regional-population/latest-release

27. Gelman A, Hill J, Vehtari A. Regression and Other Stories [Internet]. Cambridge University Press; 2020. Available from: https://users.aalto.fi/~ave/ROS.pdf

28. Australian COVID-19 Serosurveillance Network. Seroprevalence of SARS-CoV-2-specific antibodies among Australian blood donors: Round 4 update [Internet]. Kensington, NSW: UNSW Kirby Institute; 2023 Feb p. 1–9. Available from: https://www.kirby.unsw.edu.au/research/projects/serosurveillance-sars-cov-2-infection-inform-public-health-responses

29. Landis JR, Koch GG. The measurement of observer agreement for categorical data. Biometrics. 1977 Mar;33(1):159.

30. Pandemic Declaration Accountability and Oversight Committee. Review of the Pandemic (Quarantine, Isolation and Testing) Orders [Internet]. Melbourne: Parliament of Victoria; 2022. Available from: https://www.parliament.vic.gov.au/4af997/contentassets/fb24db1ca3dc41929fde603bdfca5a9a/review-of--the-pandemic-quarantine-isolation-and-testing-orders.pdf

31. Nayagam SN, Coote W, Carroll MTC, Toh ZQ, Licciardi PV, Abramson MJ, et al. SARS-CoV-2 serology and differentiation from vaccine-induced antibodies: challenges for epidemiological research. J Infect Dis. 2025;jiaf295.

