## Supplementary tables and figures for "Agreement between self-reported COVID-19 and dried blood spot serology"

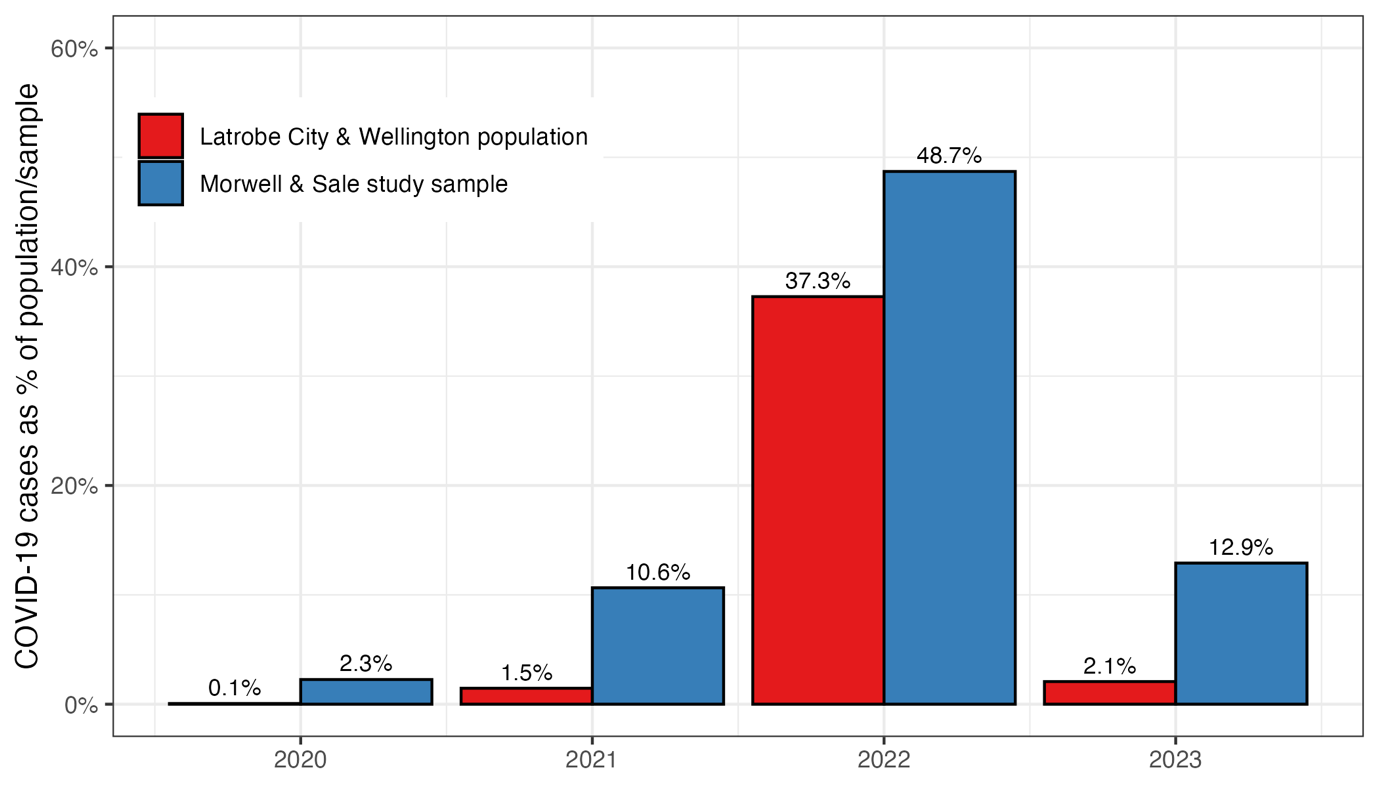
Figure S1. Annual incidence of COVID-19 in the local government area populations and annual prevalence in the study sample; self-report and serology data were collected during a 2023 clinical visit; years on the x-axis refer to retrospective answers of when infection occurred; annual incidence calculated using case data from the Victorian Department of Health (25) and Australian Bureau of Statistics population estimates for Local Government Areas (26); 2023 is a partial year both for the population data and study sample: COVID-19 cases were was no longer reported after 14 September 2023, while the study finished data collection on 6 October 2023; note that incidence (new cases per population) and prevalence (population with a case) are different but roughly comparable figures

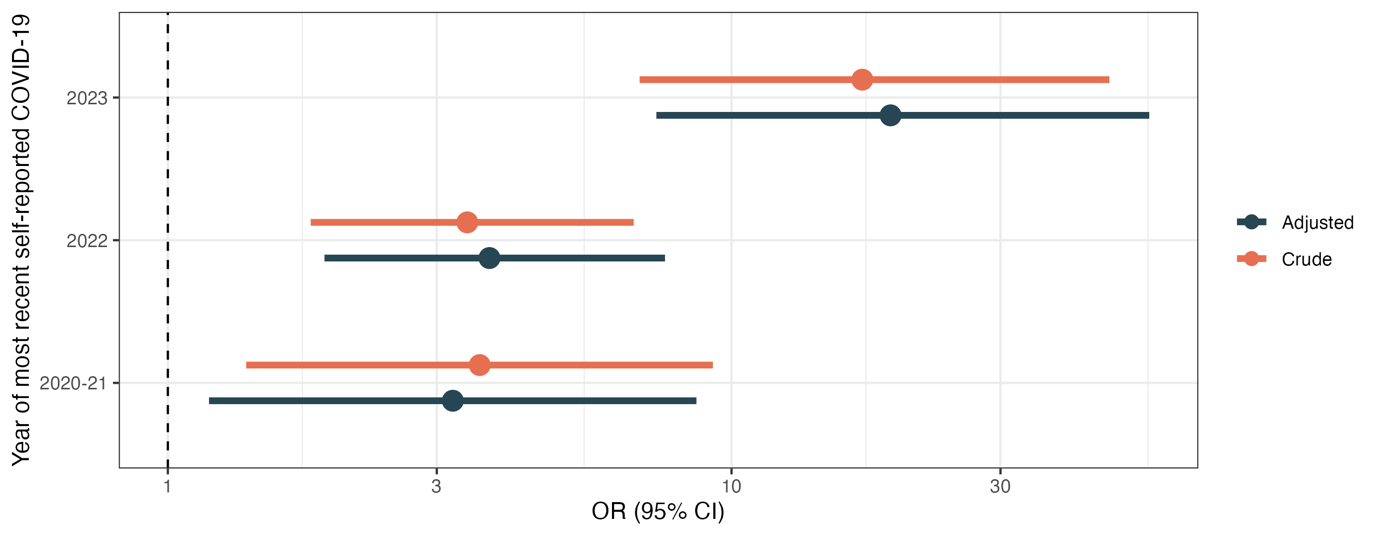
Figure S2. SENSITIVITY ANALYSIS: Crude and adjusted (age, sex, educational attainment) odds of seropositive or equivocal dried blood spot sample by year of most recent self-reported COVID-19; self-report and serology data were collected during a 2023 clinical visit; “most recent self-reported COVID-19” refers to retrospective answers of when infection occurred

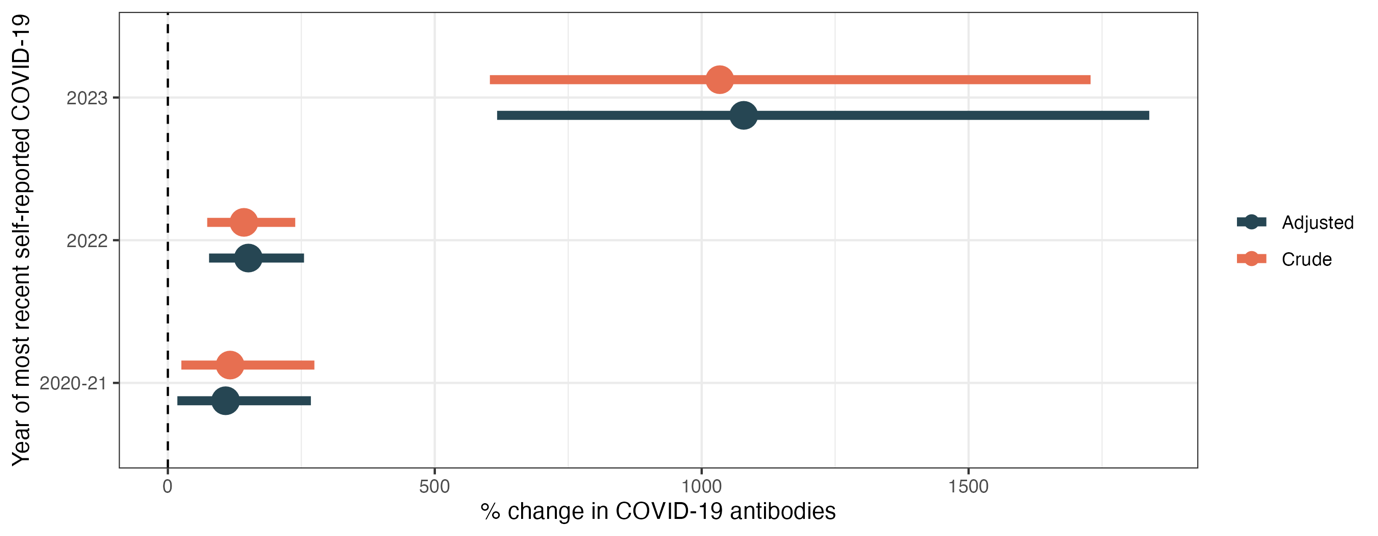
Figure S3. Crude and adjusted (age, sex, educational attainment) percent change in SARS-CoV-2 antibody N-proteins by year of most recent self-reported COVID-19; self-report and serology data were collected during a 2023 clinical visit; “most recent self-reported COVID-19” refers to retrospective answers of when infection occurred

Table S1. 2x2 table of COVID-19 cases based on self-report and serostatus, combining seropositive and equivocal serology

|  | **Seropositive** | **Seronegative** | **Totals** | **Predictive Agreement** |
| --- | --- | --- | --- | --- |
| Self-report + | 84 | 119 | **203** | 46% |
| Self-report - | 14 | 94 | **108** | 82% |
| **Totals** | **98** | **213** | **311** |  |

Note: Self-report and serological data were collected during a 2023 clinical visit; “most recent self-reported COVID-19” refers to retrospective answers of when infection occurred

Table S2. Odds of seropositivity by year of most recent self-reported COVID-19, crude and adjusted for age, sex, and educational attainment

| **Year of most recent self-reported COVID-19** | **Crude Odds Ratio** | **Adjusted Odds Ratio** |
| --- | --- | --- |
| 2020-2021 | 3.73 (1.41-9.72) | 3.50 (1.28- 9.54) |
| 2022 | 3.28 (1.72- 6.59) | 3.61 (1.84- 7.45) |
| 2023 | 20.14 (8.41-52.41) | 22.96 (9.18-62.52) |

Note: Self-report and serological data were collected during a 2023 clinical visit; “most recent self-reported COVID-19” refers to retrospective answers of when infection occurred

Table S3. SENSITIVITY ANALYSIS: Odds of seropositive *or equivocal* serology by year of most recent self-reported COVID-19, crude and adjusted for age, sex, and educational attainment

| **Year of most recent self-reported COVID-19** | **Crude Odds Ratio** | **Adjusted Odds Ratio** |
| --- | --- | --- |
| 2020-2021 | 3.03 (1.21- 7.50) | 2.84 (1.09- 7.31) |
| 2022 | 2.88 (1.59- 5.37) | 2.99 (1.62- 5.72) |
| 2023 | 16.13 (6.86-41.39) | 17.45 (7.16-46.37) |

Note: Self-report and serological data were collected during a 2023 clinical visit; “most recent self-reported COVID-19” refers to retrospective answers of when infection occurred

Table S4. Percent change in SARS-CoV-2 antibody N-proteins by year of most recent self-reported COVID-19, crude and adjusted for age, sex, and educational attainment

| **Year of most recent self-reported COVID-19** | **Crude % change** | **Adjusted % change** |
| --- | --- | --- |
| 2020-2021 | 117% (25-274) | 108% (18-268) |
| 2022 | 143% (74-239) | 151% (77-255) |
| 2023 | 1,034% (603-1729) | 1,079% (617-1,838) |

Note: Self-report and serological data were collected during a 2023 clinical visit; “most recent self-reported COVID-19” refers to retrospective answers of when infection occurred
